# Clinical Management of Cystic Echinococcosis in Bhutan: Current Practices, Gaps, and Recommendations

**DOI:** 10.64898/2026.05.08.26352730

**Authors:** Pema Wangchuk, Jan Hattendorf, Jakob Zinsstag, Thomas Junghanns

## Abstract

**Background:** Cystic echinococcosis (CE) is a neglected parasitic zoonosis that primarily affects marginalized populations in rural endemic regions. These populations have limited access to, and availability of, the appropriate infrastructure, resources, and skills required to treat this complex disease. This retrospective study reviewed and analyzed the clinical management of CE patients in Bhutan, based on hospital records from January 2020 to December 2024.

**Methods:** Hospital records of 120 patients with hepatic or pulmonary CE treated between January 2020 and December 2024 in the three hospitals caring for CE patients in Bhutan were retrospectively reviewed. Data on clinical presentation, diagnosis, treatment, and outcomes were extracted from hospital records using a standardized questionnaire. Data analysis was performed using R software (version 4.4.3).

**Findings:** The median age of the patients was 36 years (IQR: 21.75–53), with 60% being female. The liver was the most affected organ (70%), followed by the lungs (8%). US and CT were used in 83% for diagnostic and pre-surgical assessments. WHO-recommended CE cyst staging was performed in only 11% of cases. Pre-intervention complications were reported in 40% of patients. Treatment approaches included surgery (open or laparoscopic partial cystectomy, deroofing, and cyst drainage) combined with albendazole (ABZ) therapy (74%), PAIR in 4%, and ABZ alone in 8% of cases. Antibiotic use beyond standard perioperative prophylaxis was common (44% of cases). Post-treatment complications occurred in 23% of surgical cases, including one death; biliary leakage was the most frequent complication (55%), and more than one-third of surgical patients were discharged with drains in situ. 23% of the cohort were readmissions and 11% of the patients with hepatic CE were due to documented recurrence requiring repeat surgery. Long-term follow-up was absent, limiting the early detection and management of recurrence.

**Conclusion and recommendations:** The study findings show that the care for CE patients in Bhutan urgently requires implementation of US-based cyst staging and treatment allocation, the development of infrastructure and skills for the major treatment modalities recommended by WHO guidelines and long-term follow-up to improve patient outcomes, including recurrence, and to ensure quality control of CE care. Safe and proven practices, particularly in surgery, must be prioritized over diversification. Such strategies are feasible and cost-effective.

**Author Summary:** Cystic echinococcosis (CE) is a neglected parasitic disease that develops silently and can affect people for years before causing serious complications. It is common in rural, livestock-rearing regions, including Bhutan, where access and availability to appropriate care is limited. We reviewed hospital records of 120 patients treated for CE in Bhutan between 2020 and 2024 to understand current clinical practices and treatment outcomes. Most patients had large liver cysts and were diagnosed at a late stage, often only once complications had occurred.

US-based cyst staging was very rarely performed, and cyst-staging was not used to inform treatment decisions. Surgery combined with anti-parasitic (albendazole) therapy was the most common treatment. Postoperative complications and disease recurrence were frequent, and most patients had no long-term follow-up to attend complications and recurrences timely. Considerable variation in surgical and medical management was observed. Developing and implementing WHO-guideline-based infrastructure, resources and training for CE patient care is urgently needed.

## 1. Introduction

Human cystic echinococcosis (CE) is a zoonotic parasitic disease caused by the larvae (metacestodes) of *Echinococcus granulosus* sensu lato (s.l.) tapeworm [1].

Human CE has a long incubation period; more than 10 years is not unusual. All organs can be affected; most commonly the liver, followed by the lungs. Symptoms are mostly nonspecific and include upper abdominal discomfort and cough. CE rarely presents with complications, such as cyst rupture, secondary bacterial infection, and compression syndromes. In a substantial proportion of patients, CE transitions from active to inactive cyst stages unnoticed without the need for intervention [2].

CE is diagnosed and staged using imaging, mainly ultrasound (US), which is also increasingly available and affordable in low-resource settings. Magnetic resonance imaging (MRI) widely reproduces US-defined imaging features; computed tomography (CT) does so to a much lesser extent [3]. The four main treatment modalities are drug therapy with benzimidazoles, preferably albendazole (ABZ), percutaneous cyst sterilizing methods and surgery combined with ABZ, and ‘watch and wait’. The choice is based on the cyst stage, size, location, presence of complications (e.g., cysto-biliary and cyst-bronchial fistulas), and is tuned to the healthcare infrastructure, availability of materials, and health personnel training [4,5]. Long-term follow-up of patients with CE is important to identify recurrences early, which is the main problem in CE management [6]. A five-year follow-up period has been widely recommended [2].

Little information is available on the clinical management and patient outcome in the southern Himalayas, where Bhutan is located. To fill this gap, the main objective of this study was to describe the current CE management in Bhutan to inform clinical practice and health policy.

## 2. Materials and methods

This study retrospectively reviewed the medical records of patients with CE. Medical record technicians from all referral and district hospitals, where surgical management of CE is performed, were contacted to screen and extract data. In addition to cases (n = 111) from the Jigme Dorji Wangchuck National Referral Hospital, five cases were included from the Eastern Regional Referral Hospital in Mongar, and four cases from Wangdicholing Hospital in Bumthang. The medical records of all patients with CE (ICD code B67) admitted to these hospitals between January 2020 and December 2024 were reviewed. Patients admitted for other conditions but with CE as the second diagnosis were also included in the medical records review. Information on sociodemographic factors, clinical presentation, diagnosis, and treatment was extracted using a pre-formed and pilot-tested questionnaire developed on the Open Data Kit (ODK) data collection platform. From mid-2024 onwards, imaging (CT and US), and histopathological reports not consistently stored in the paper-based records, were additionally reviewed in the electronic patient information system (ePiS).

Ethical approval for the study was granted by the Research Ethics Board for Health (approval number: Ref.No./PO/RL/2024.1.NNW), and the study protocol was additionally reviewed by the Ethikkommission Nordwest- und Zentralschweiz, Switzerland (Study ID: *AO_2024_00045*).

### 2.1 Statistical Analysis

Descriptive statistics, such as proportions, were used to describe the distribution of CE cases according to sociodemographic and clinical information. Demographic information, including age, sex, history of previous admission for CE, and clinical information, including symptoms and signs on admission, was summarized. The diagnostic method, anatomical location, treatment method, complications, antiparasitic medication, and treatment outcomes were summarized. Numerical data are described using the median and interquartile range (IQR).

## 3. Results

A total of 120 patients were diagnosed with CE as primary or secondary diagnosis in the hospitals which treat CE surgically in Bhutan between 2020 and 2024. 27 of the 120 CE cases (23%) were readmissions. Among the ‘only hepatic’ CE cases (n = 84), 20 patients were readmitted; of whom 9 (11%) were due to documented CE recurrence requiring repeat surgery. In contrast, among the ‘only pulmonary’ CE cases (n = 10), seven (70%) were readmitted none for recurrence. Four patients had been readmitted due to postoperative complications following thoracotomy, while three were admitted for further lung cyst evaluation or management of co-morbidities.

Table 1 shows the age and sex distribution of the retrospective cohort on patients treated surgically for CE.

**Table 1:** Demographic Characteristics of patients with CE.

| Category | Count (%) |
| --- | --- |
| **Age groups** |  |
| Children (0 – 14 years) | 15 (12.5%) |
| Adults (15 to 64 years) | 91 (75.8%) |
| Elderly (65 + years) | 14 (11.7%) |
| **Sex** |  |
| Male | 48 (40%) |
| Female | 72 (60%) |

Of the 120 patients, 108 (90%) had CE as the primary diagnosis. Among them, 80 patients (74%) had reported at least one symptom upon hospital admission. The most common symptom was abdominal pain (right hypochondrial pain, epigastric pain, and abdominal discomfort). The details of symptoms at hospital admission by CE cyst organ involvement are presented in Figure 1. 48 (44%) patients had positive clinical signs on examinations at admission. Among those, right hypochondrial (RHC) tenderness was the most common finding, reported in 34 (70%) patients.

**Figure 1:**
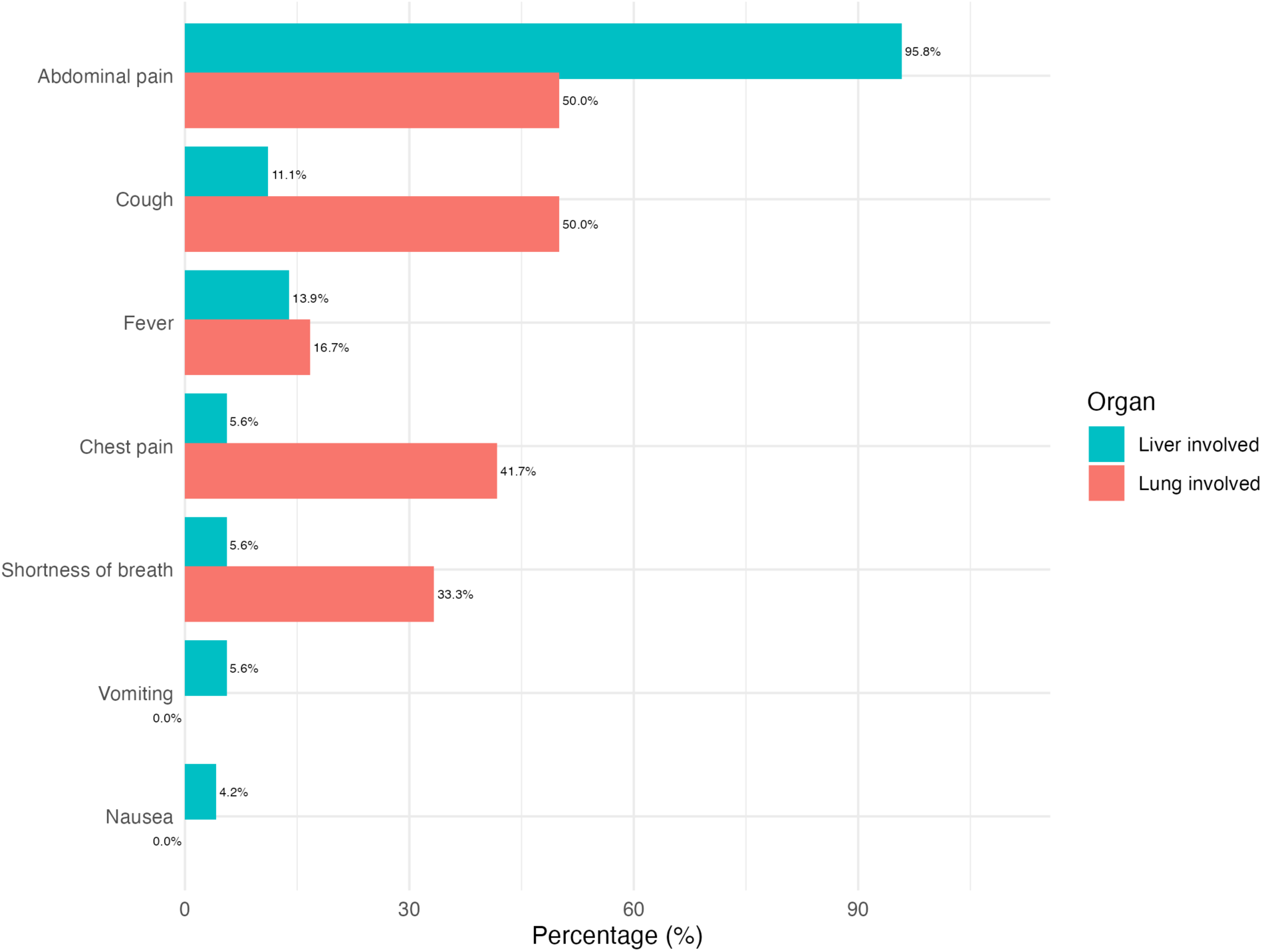
Frequency and symptom types based on CE organ involvement.

Table 2 shows the distribution of CE cysts within and between organs. The maximum number of cysts per patient was five, with an average of 1.5 cysts per patient. Cyst size was recorded in 88 patients (73%). The median cyst size was 10 cm, with an interquartile range of 7.9 – 14.1 cm. The minimum cyst size was 5 cm, with a maximum of 22.7 cm.

**Table 2.** Distribution of CE cases by Organ Involvement.

| Location | Number of Patients | Percentage (%) |
| --- | --- | --- |
| Only liver | 84 | 70% |
| Liver & other location | 13 | 10.8% |
| Only lung | 10 | 8.3% |
| Both liver & lung | 8 | 6.7% |
| Other rarer locations* | 3 | 2.5% |
| Lung & other location | 2 | 1.7% |

\* *retroperitoneum (2) & pleura (1)*

In all cases, diagnoses were made based on imaging findings. The most commonly used imaging methods were US, CT, X-ray (chest radiography) (Figure 2). Serological tests (ELISA) were used as complementary tests in only nine (8%) cases, of which five (56%) were positive and four (44%) were negative in surgically and or histopathologically confirmed CE patients. Histopathological confirmation reports were available for 89 (74%) of the 120 patients with CE.

**Figure 2.**
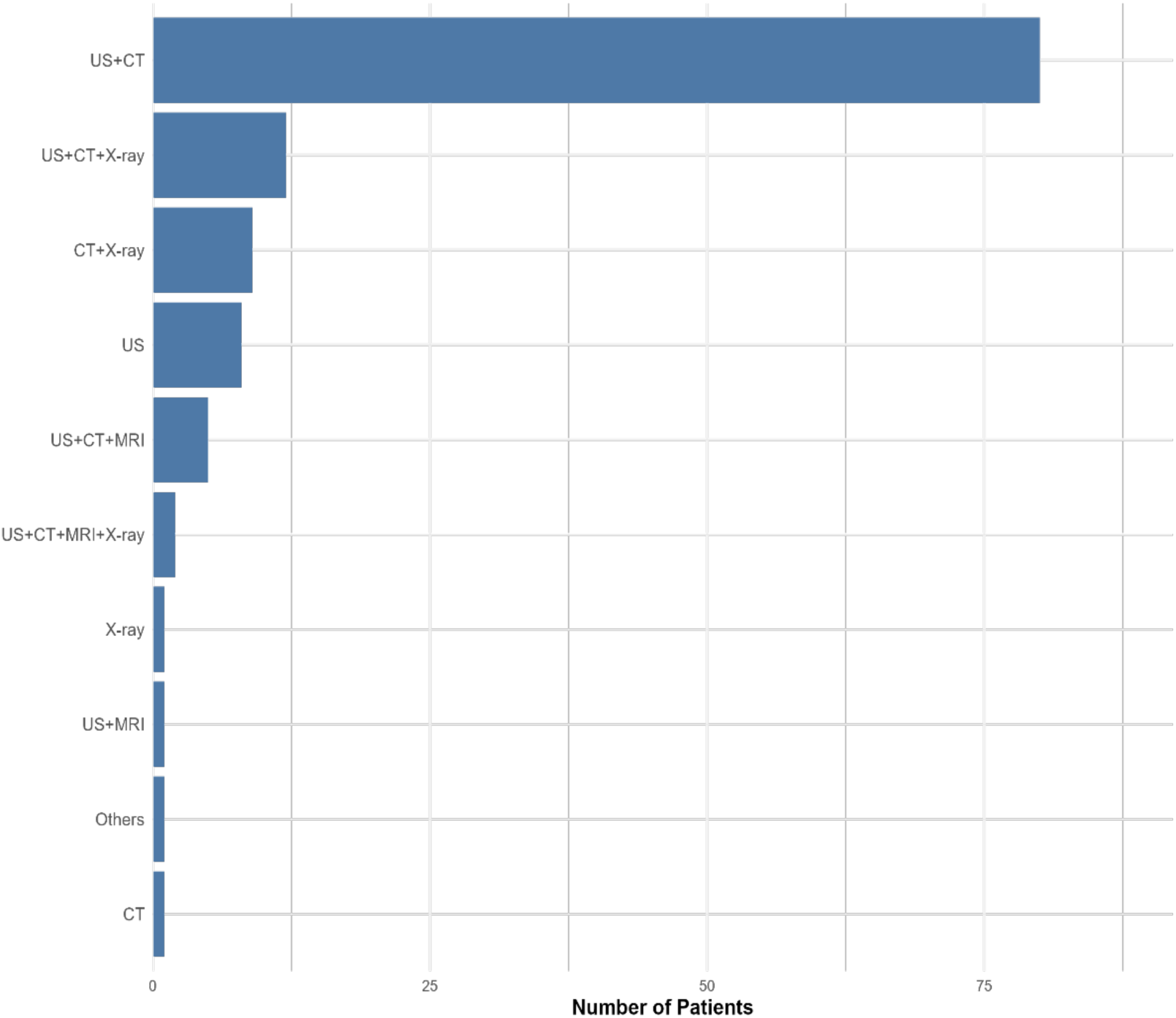
Combination of imaging methods used for CE diagnosis.

CE staging was performed in only 13 (11%) cases. The WHO CE cyst staging classification was applied in five cases: two cysts were classified as CE3a, and one each as CE3b, CE1, and CE2; the Gharbi classification was used in six cases; three were Gharbi type II, two Gharbi type I, and one Gharbi type III. Both classifications were applied in one case (WHO stage CE3a and Gharbi Type II). Of the 120 cases, operative (surgeons’) notes were available for 104 patients (87%). However, detailed cyst-specific intraoperative findings were documented only in a subset of these records. From 104 available operative notes, daughter cysts were reported in 31 cases (31/104, 26%), membranes in six cases (6/104, 6%), and membrane and daughter cysts in four cases (4/104, 3%). Two patients (2/104, 2%) had a solid cyst. In the remaining operative records, cyst-specific findings were not explicitly documented.

Pre-intervention cyst complications were reported in 48 patients (40%). Most patients (35/48, 73%) had a single complication, 13 (27%) had more than one complication. A total of 54 complications were observed in 48 patients. Out of 54 different complications, 23 (43%) events were infections or features suggestive of infection, nine (17%) had biliary fistulae, seven (13%) features of compression syndrome, five (9%) cyst rupture, one (2%) venous/arterial complications, five impending cyst rupture, one patient had intra-abdominal and pelvic, one intra-thoracic, and one epigastric CE manifestations, one empyema thoracis, and nine (17%) patients ‘other complications’.

Treatment modalities are reported by affected organs. Patients with concomitant hepatic and pulmonary CE are, therefore, represented in more than one treatment category. In addition, two cases did not receive CE-specific treatment at the time of the study. The most common treatment was surgery combined with anti-parasitic (ABZ) therapy (89/120, 74%). For hepatic and pulmonary CE, the treatment combinations are shown in Tables 3A and 3B, stratified by cyst size. Almost one-third of the patients (28%, 34/120) received antibiotics; 15 patients (44%, 15/34) of those at the time of admission due to clinically suspected infected cyst (microbiological confirmation was unavailable); 47% (16/34) pre-operative prophylaxis. In two patients (6%, 2/34), antibiotics were started post-operatively due to complications (pneumonia and paralytic ileus). In one patient (3%, 1/34) with hepatic CE, the patient was treated for community-acquired pneumonia.

**Table 3A.**
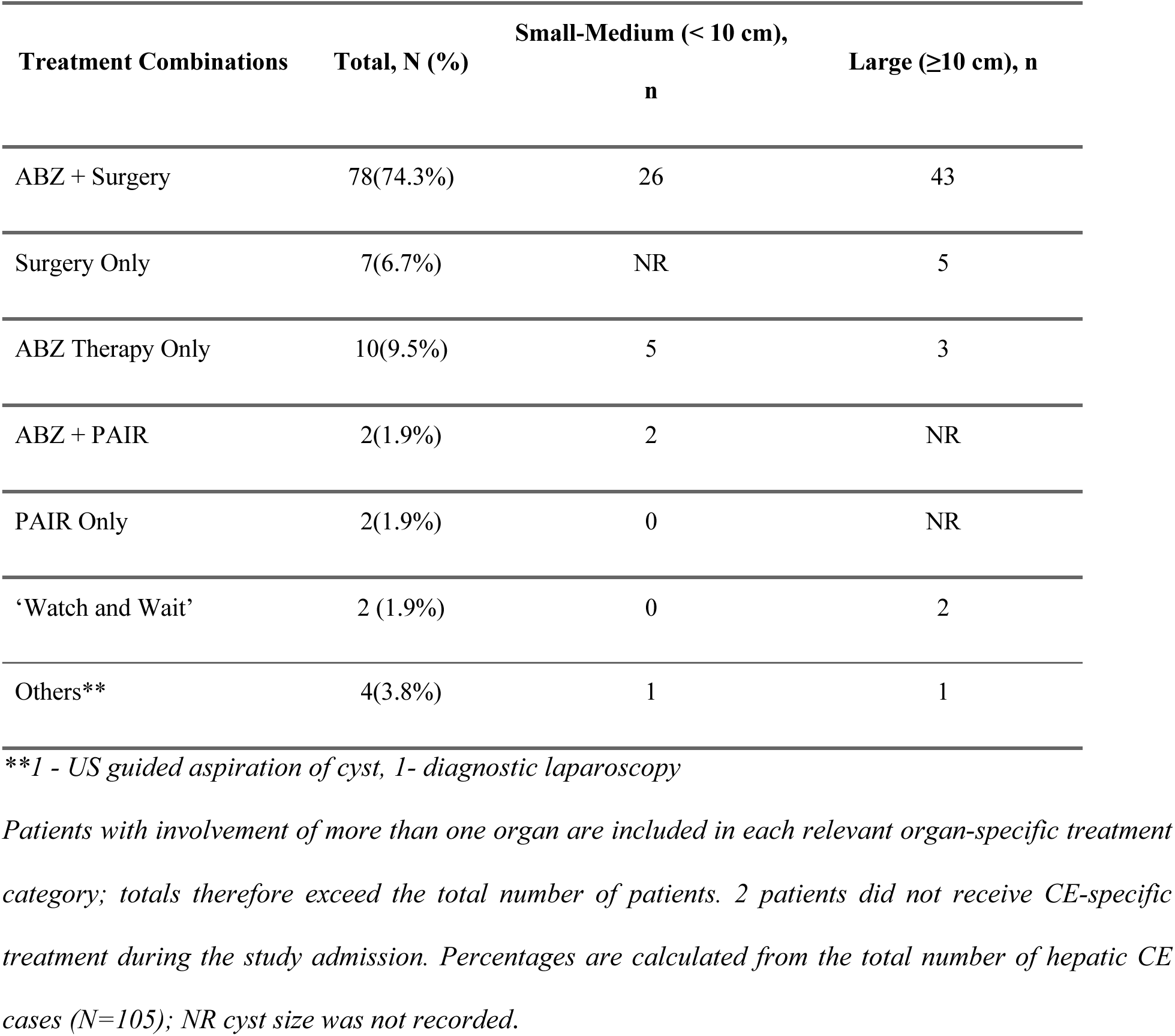
Treatment for hepatic CE by cyst size.

**Table 3B.**
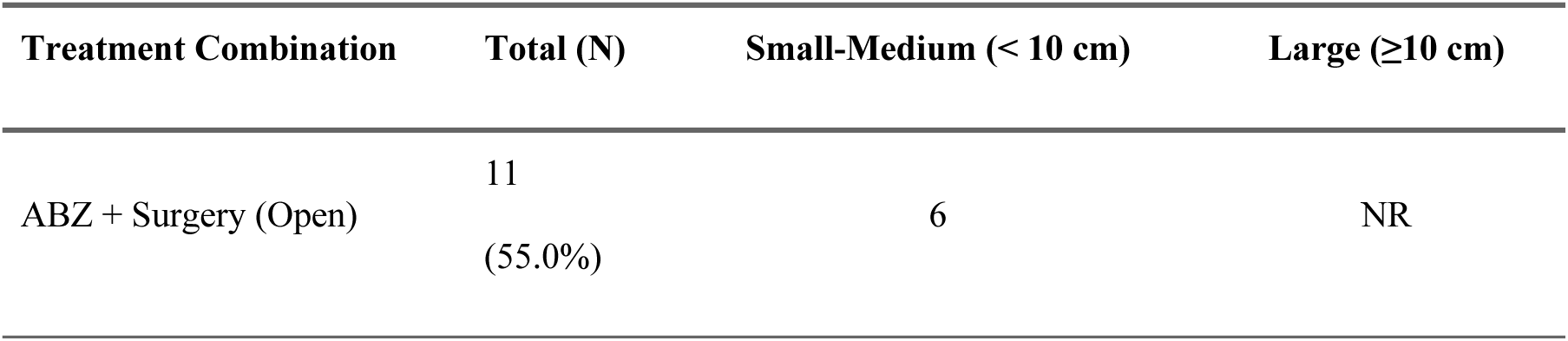
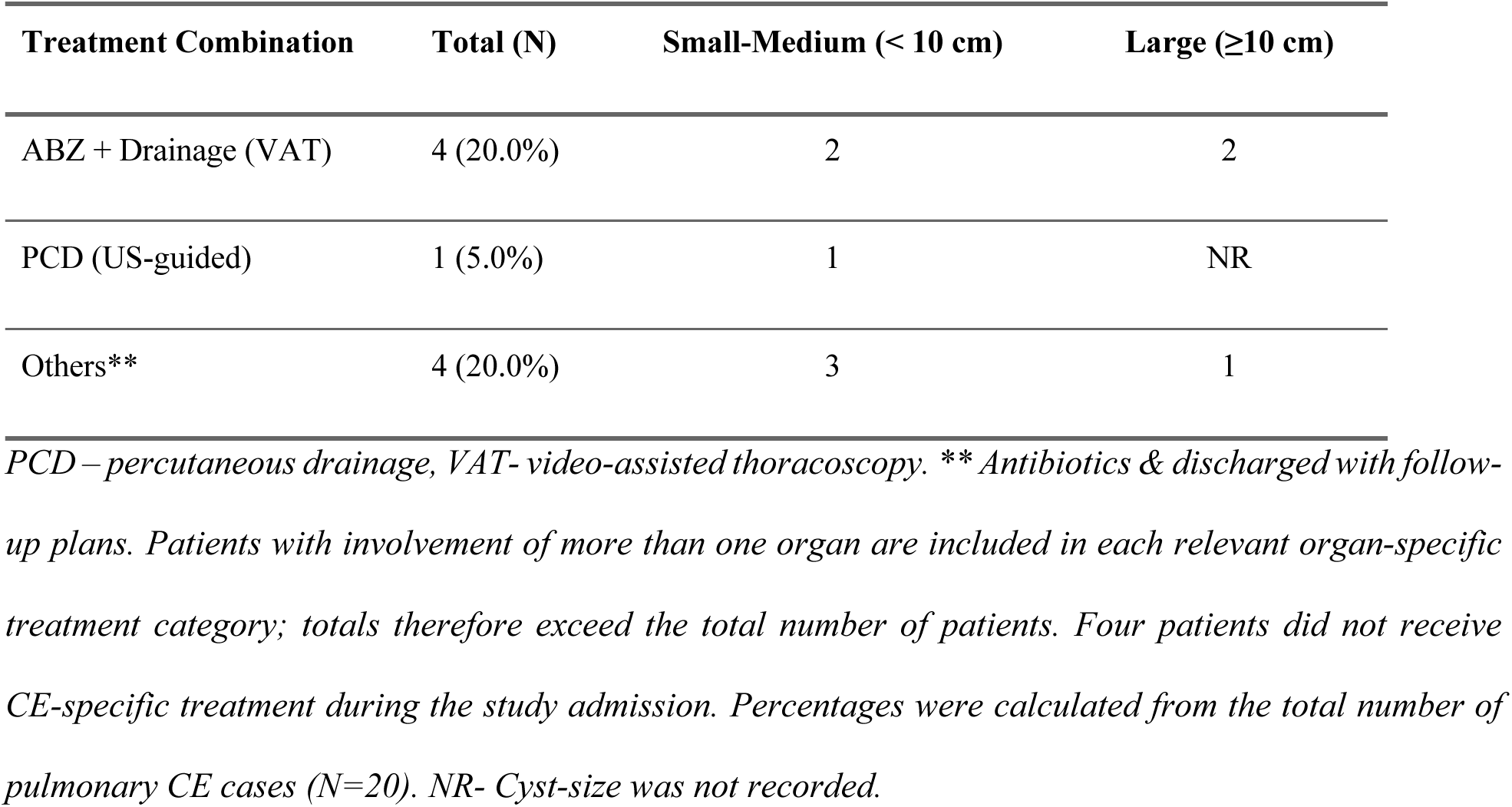
Treatment for pulmonary CE by cyst size.

Of the 96 patients who underwent surgery, laparoscopic deroofing and cyst drainage was attempted in 59/96 (61%), completed successfully in 52 (54%) and converted in 7 (7%); primary open surgery was performed in 30/96 (31%). Among the seven (7%; 7/96) surgically treated pulmonary CE cases, five underwent video-assisted thoracoscopic (VAT) cyst deroofing and drainage, one open thoracotomy and drainage of the pleural cyst and ICT drainage, and one ICT drainage and chemical pleurodesis.

In terms of spillage prophylaxis, in addition to preoperative albendazole administration, surgical fields were covered with protoscolecidal-soaked gauze in all cases (100%, 96/96). Povidone iodine (10 %) was used as a protoscolecidal agent in 95 cases, and 20% hypertonic saline in one case.

Postoperative complications were reported in 22 of 96 (23%) patients. The most common post-surgical complication was biliary leakage (ICD code K91.89), defined as post-operative bile leak from the drain, which was observed in 12 of 22 cases (55%). One-third of the patients (4 of 12, 33%) who developed post-surgical biliary leakage had a cysto-biliary fistula preoperatively. All patients with postsurgical biliary leakage were conservatively managed with daily drain output monitoring. In one case, leakage stopped spontaneously during the hospital stay, whereas 11 patients (92%) were discharged with a drain and a follow-up plan within 1 week after discharge. In all cases, the drain output showed a decreasing trend at the time of discharge from the hospital. A detailed breakdown of the post-surgical complications is presented in Table 4. Most patients stayed in the hospital for < 10 days, with a median of 5 days (IQR: 3 – 8 days).

**Table 4.**
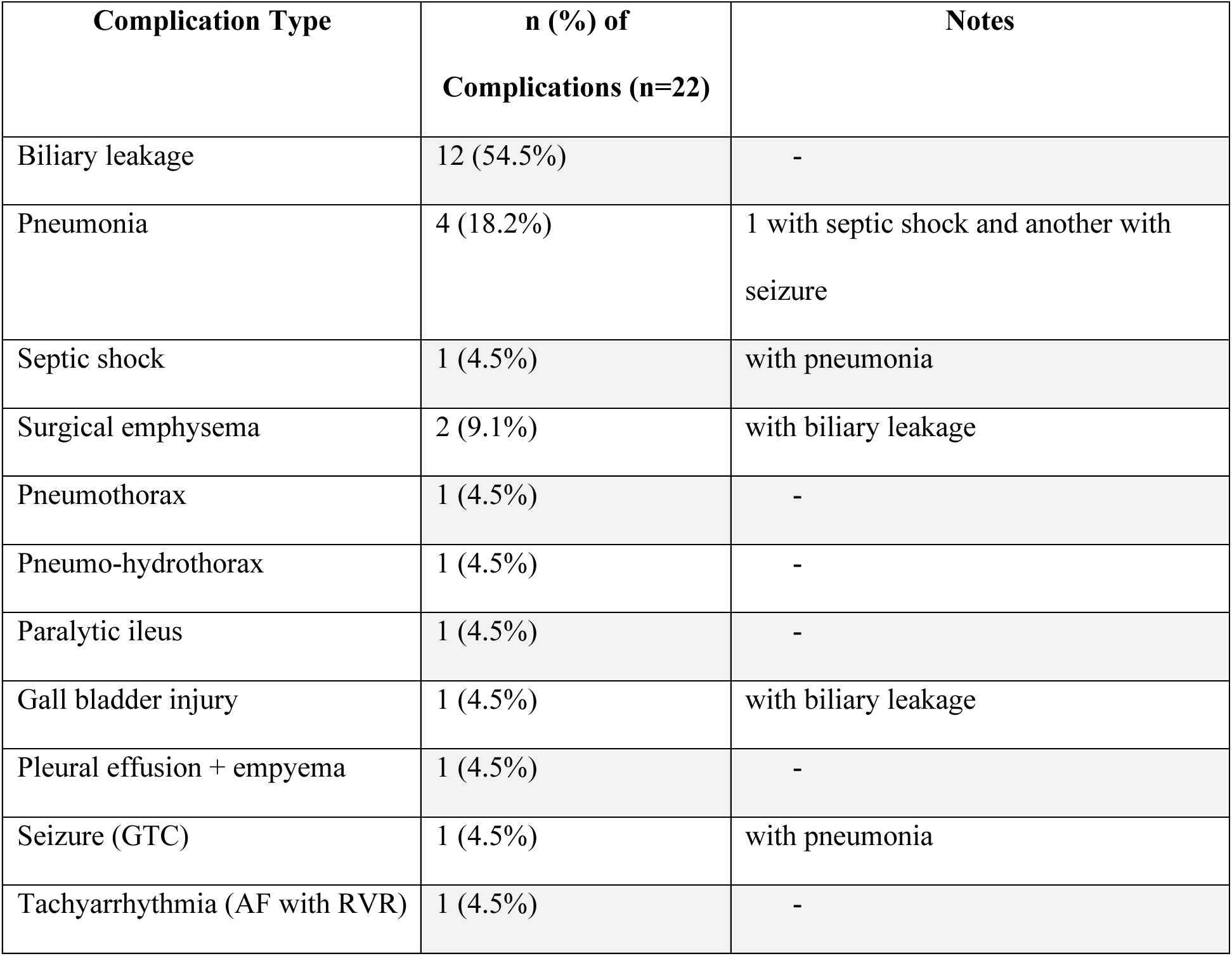
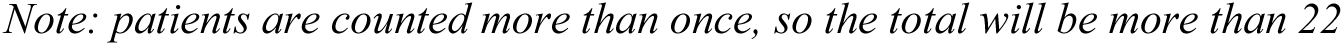
Reported postoperative complications in patients with CE.

| Complication Type | n (%) of<br>Complications (n=22) | Notes |
| --- | --- | --- |
| Biliary leakage | 12 (54.5%) | - |
| Pneumonia | 4 (18.2%) | 1 with septic shock and another with seizure |
| Septic shock | 1 (4.5%) | with pneumonia |
| Surgical emphysema | 2 (9.1%) | with biliary leakage |
| Pneumothorax | 1 (4.5%) | - |
| Pneumo-hydrothorax | 1 (4.5%) | - |
| Paralytic ileus | 1 (4.5%) | - |
| Gall bladder injury | 1 (4.5%) | with biliary leakage |
| Pleural effusion + empyema | 1 (4.5%) | - |
| Seizure (GTC) | 1 (4.5%) | with pneumonia |
| Tachyarrhythmia (AF with RVR) | 1 (4.5%) | - |

*Note: patients are counted more than once, so the total will be more than 22*

Albendazole was the only prescribed anti-parasitic medication. Sixty-three (63/120, 53%) patients received albendazole therapy during hospital stay. The duration of pre-admission albendazole therapy could not be determined. Upon discharge from the hospital, 96 (80%) patients were treated with albendazole. Regarding post-hospitalization albendazole therapy, the duration of therapy was known in 90 (75%) patients. Of those, 85 (94%) received continuous therapy (without breaks in between), and the median duration of therapy was 1 month (IQR: 1–2). In the remaining five patients (5/90,6%), the median therapy duration was 1 month (IQR, 1–3). They received interrupted albendazole therapy (1 week break after every cycle, that is, four weeks of therapy).

In terms of discharge and post-discharge outcomes, one patient died three months postoperatively due to pneumonia and septic shock (1/120, 1%), one patient (1%) had early postoperative morbidity (persistent pus drainage), one patient (1%) was referred to a higher center for further management, one patient was discharged with an MRCP plan, six patients (5%) were discharged with a surgical follow-up plan, 43 of 96 surgical patients (inclusive of 11 with biliary drains) (45%) were discharged with drains.

## 4. Discussion

This hospital-based retrospective review of the medical records of patients with CE admitted for treatment between January 2020 and December 2024 provides a comprehensive clinical analysis of the hospital management of CE patients in Bhutan over a five-year period. It includes CE diagnostics, mainly imaging, clinical presentations, CE organ involvement, complications (pre-and post-treatment), and the treatment approaches used.

### 4.1 Demographic patterns

The predominance of CE cases among women (60%) is consistent with the findings of previous hospital-based studies in Bhutan and other endemic regions [7,8]. The predominance of CE cases among adults aged 15–64 years (76%) reflects the long incubation period of the disease and transmission pressure, with clinical manifestations often appearing years after the initial infection [9]. At the same time, this finding should be interpreted in the context of Bhutan’s demographic structure, where this age group represents the largest demographic segment [10].

### 4.2 Clinical presentation and organ distribution

The liver was the most frequently affected organ, which is consistent with regional and global patterns.

Most patients in this cohort presented with symptoms and signs on admission, which were non-specific and mild, as it is reported in other studies. The most common clinical presentation is related to the abdomen, as the majority of cases had liver involvement. It is important to note that large cyst size at presentation in this study indicates late detection and prolonged asymptomatic disease progression or health services access barriers. From a clinical and public health perspective, such findings show the importance of early detection at primary healthcare level, before cysts reach sizes associated with higher risk of complications.

### 4.3 Diagnostics and CE cyst staging

An important finding of this study was the limited use of WHO-recommended CE cyst staging. Although US and CT were widely used for diagnosis and presurgical assessment, cyst staging was performed in only 11% of cases. The WHO CE cyst classification is recognized for guiding treatment decisions, monitoring treatment response and detecting recurrence during follow-up. The guidelines recommend the use of US-based CE cyst staging to guide allocation to the appropriate treatment modality [4,11]. The underutilization of staging in this cohort might be due to a lack of awareness for and training in CE cyst staging of sonographers and clinicians, as well as the lack of standardized protocols/guidelines to guide CE treatment in the country.

In the absence of pre-operative US-based staging, operative findings provide indirect insight into the cysts selected for surgical treatment. Among the 120 cases, surgical notes were available for 87% of the patients, of whom approximately 39% had detailed cyst-specific intraoperative findings consistent with active disease, including the presence of daughter cysts only (26%), laminated membrane (6%) or both (3%). These morphological features of cysts are well-established indicators of cyst viability and active CE cyst stages [4]. The findings of the retrospective review of medical records cannot, however, establish the appropriateness of surgical indications at the individual patient level since preoperative CE cyst staging was performed in only a minority of patients and intraoperative and histopathological information was available in only a subset of operated patients, limited to fewer than half. Additionally, a high proportion of patients in the cohort were admitted for complications. Overall, overtreatment of CE is, however, likely, when preoperative CE cyst staging is not done. In CE surveys in endemic countries 50% of inactive cysts are not rarely found [12]. Therefore, implementation of CE cyst staging is necessary to enable stage-specific treatment decision, optimizing treatment outcomes, including the reduction of unnecessary high-risk surgeries for inactive cysts. Additionally, disease reporting for clinical and epidemiological surveillance is improved.

Imaging modalities, particularly US, constitute the primary means of diagnosing CE in this study; often combined with CT. The latter is not a primary diagnostic imaging modality for CE since it is weak on staging as compared with MRI as an additional tool for diagnosing and staging CE [3]. This reflects global best practices in which US is the first-line diagnostic approach for CE [13], and CT is a useful tool for presurgical planning.

Serology was used in only a small proportion of patients in this cohort (8%). Current evidence and WHO-IWGE emphasize that CE diagnosis should primarily be based on imaging. Among the few patients tested, ELISA was negative in almost half of the patients tested despite imaging and surgically confirmed CE. Thes findings reinforce that negative serological results do not exclude CE. Strengthening imaging-based diagnosis and routine cyst-staging therefore remains the most relevant, and cost-effective diagnostic strategy.

### 4.4 Treatment practices

Our study findings showed that most patients with CE (56%) received a combination of different treatment modalities, of which surgery combined with albendazole was in line with WHO guidelines recommendations for appropriate CE cyst stages [4,11]. Antiparasitic therapy was administered to 80% of discharged patients, an important perioperative adjunct to surgery. The observed treatment duration and the use of interrupted courses are, however, inconsistent with WHO-recommended regimens.

From a clinical management perspective, effective CE treatment depends on the consistent implementation of well-established, stage-specific strategies, guided by US-based cyst staging. The strategies need to be tried and tested and adapted to the given infrastructure, resources, and skills [2,4,11]. The relatively high proportion of laparoscopic approaches, including video-assisted thoracoscopic (VAT) procedures for pulmonary CE (60%) shows the increasing adoption of minimally invasive procedures in the country. While use of laparoscopic surgery in selected uncomplicated cases may be considered [11], and is increasingly used [16], claims of benefit are often based on short-term post-operative surrogate rather than long-term outcomes, most importantly disease recurrence. Recurrence after surgical treatment is the most important disease-specific outcome in CE management, however, implementation of well-established measures to prevent recurrence remains low. The 7% conversion rate from laparoscopic to open surgery in our study shows the importance of appropriate patient selection and a realistic assessment of surgical expertise.

Percutaneous methods, particularly PAIR, were used in only 3% of cases. Although there is a lack of comparative studies (surgery versus PAIR, in combination with drugs), PAIR, as a less invasive treatment options in CE management appears to be non-inferior to surgery in terms of recurrence and postoperative complications in situations where there is no biliary communication [11]. The limited use of PAIR in the three major hospitals treating CE in Bhutan suggests the need for broader dissemination and training of WHO-recommended less-invasive techniques. Clinical common sense stresses that diversification into advanced surgical approaches, including laparoscopic techniques, should be deferred until conventional surgery, PAIR, medical therapy, and watch-and-wait strategies are fully established, standardized, and supported by adequate training and follow-up [2,4,11].

Protoscolecidal agents used in this setting included 10% povidone iodine as the primary agent and 20% hypertonic saline used in one case. 20% hypertonic saline is generally recommended including a WHO expert consensus (4). Povidone iodine is a weak protoscolecidal agent and has not been endorsed by WHO. The use of protoscolecidal agents is contraindicated when cysto-biliary communication is suspected or confirmed [11].

### 4.5 Complications, outcomes and follow-up

Our study shows that pre-intervention CE cyst complications were common, with close to half of the cases (40%) reporting at least one complication. Around 13% were recorded as having suspected cyst infection. Although rare, secondary bacterial infection of liver echinococcal cysts occur [17,18]. Infection rates are higher in complex cystic lesions [18]. The high proportion of antibiotic use outside of standard perioperative prophylaxis observed in this cohort, is difficult to interpret in the absence of appropriate microbiological work-up and/or documentation of results in the medical records. Expert consensus and the WHO-IWGE aligned reviews emphasize that secondary bacterial infection of CE cysts is uncommon and frequently over-suspected, leading to unnecessary antibiotic exposure in the absence of standardized diagnostic and treatment algorithms [2,18].

The presence of biliary fistulae (15%) in our study is similar to that reported elsewhere [16]. Biliary fistulae increase the risk of postoperative complications. Identification and safe closure must be achieved. Other pre-intervention complications observed in our cohort include compression syndrome (11%) and cyst rupture (8.1%) indicating late clinical presentation and show why timely diagnosis and intervention matter [2]. Across the cohort, the postoperative complication rate of (23%) is within the range reported in the literature. Among postoperative complications, biliary leakage constituted 54.5% of all postoperative complications [16], as reported in recent series and reviews of CE management.

Short-term outcomes in this cohort were not satisfactory, despite reported clinical improvement in a proportion of patients which, however, could not be specified based on the data extractable from the medical records. More than one-third were discharged with surgical drains in situ. Discharging patients with surgical drains in-situ is not a standard practice and reflects unresolved postoperative complications, most commonly biliary leakage. Close monitoring of patients for potential complications after surgical interventions is recommended [11]. Persistent external drainage should prompt inpatient evaluation and definitive management including imaging and appropriate interventions rather than outpatient observation. Prolonged biliary drainage is associated with increased morbidity, infection risk, and delayed recovery. This practice is particularly problematic in resource-limited settings with weak follow-up systems, where loss to follow-up may delay recognition and treatment of complications, thereby increasing the risk of avoidable short- and long-term harm.

Documented recurrence requiring repeat surgery occurred in 9 of 84 patients (11%) with hepatic CE. Postoperative recurrence remains one of the most important clinical outcomes in CE management, since it drives repeat hospitalization, additional interventions, and prolonged morbidity, with pooled global recurrence rates after primary resection ranging widely across regions and surgical techniques. The 2025 WHO guidelines for the treatment of patients with CE recommend US-based cyst staging combined with stage-specific treatment as the principal strategy to reduce recurrence and standardize care [11]. Long-term, structured follow-up was not available in this study; therefore, the 11% figure reflects only those patients who returned to the same hospitals for re-operation and almost certainly underestimates the true burden of recurrence.

### 4.6 Strengths and Limitations of the Study

The study population is likely to capture the vast majority of treated CE cases in the country supporting the representativeness of the findings at the national level since CE is treated in only the three hospitals covered by the study. The Jigme Dorji Wangchuck National Referral Hospital serves as the country’s main tertiary referral center and is the only hospital where sub-specialties for hepatic and cardio-thoracic surgeries are located. The Eastern Regional Referral Hospital serves as the main tertiary referral center for the entire eastern region, and Wangdicholing Hospital in the central region serves as the regional cluster hospital where surgical services are available. Together, these institutions constitute the core national infrastructure for CE management, and patients requiring diagnostic confirmation, surgical intervention, or management of complicated CE are routinely referred to these centers particularly the national referral hospital in Thimphu.

However, the completeness and quality of the data were influenced by reliance on paper-based medical records for much of the study period. Until the recent rollout of ePiS in the country, imaging reports, particularly US images are handed over to patients at hospital discharge, and copies of detailed operative and follow-up documentation, were not consistently archived affecting the quality of retrospective review exercises, such as in this study. This limited the retrospective assessment of CE cyst staging and longitudinal outcomes. Consequently, retrospective CE cyst staging using the WHO-IWGE US classification was not feasible. Strengthening imaging data management, including US images and videos, is essential for improving diagnostic and therapeutic capacity and longitudinal patient follow-up.

Owing to the lack of systematic follow-up of patients and the difficulty in reaching out to patients after hospital discharge, clinical outcomes, including long-term complications and recurrence, cannot be assessed. It is not unique to Bhutan and has been reported from other endemic regions [1,9]. The recent rollout of the ePiS system represents an important step toward improving data completeness, standardized documentation, imaging archiving, and longitudinal follow-up. Even for the current study, the ePiS platform enabled access to more complete clinical information that had not been routinely archived in earlier years. This improvement was most evident in cases managed in late 2023 and 2024.

## 5. Conclusion & Recommendations

The study findings show that the care for CE patients in Bhutan urgently requires implementation of US-based cyst staging and treatment allocation, the development of infrastructure and skills for the major treatment modalities recommended by WHO guidelines and long-term follow-up to improve patient outcomes, including recurrence, and to ensure quality control of CE care. Safe and proven practices, particularly in surgery, must be prioritized over diversification. Such strategies are feasible and cost-effective.

## Acknowledgments

We thank the medical records technicians from the Jigme Dorji National Referral Hospital in Thimphu, Eastern Regional Referral Hospital in Mongar and Wangdicholing Hospital in Bumthang for their assistance in reviewing and extracting relevant data from the medical records. We thank Tshewang Dema, Chimi Thinley, Laxmi Prasad (clinical nurses) of Gidakom Hospital, and health staff (sonographers) and clinicians of regional and district hospitals (Phuntsholing, Gedu, Bumthang, Trongsa, Bajo, Punakha, Paro, Haa, Chukha) for their support.

## Data Availability Statement

All relevant data are within the manuscript. The anonymized dataset is available from the corresponding author upon a reasonable request. In addition, the Ministry of Health owns the full dataset. Interested individuals may obtain the data upon request from the Chief Planning Officer, Policy and Planning Division, Ministry of Health, P.O. Box: 726, Kawangjangsa, Thimphu, Bhutan; Phone number- +975-2-328095, 321842.

